# Mortality from Sickle Cell Disease in Brazil

**DOI:** 10.1101/2023.05.26.23290600

**Authors:** PF Blatyta, C DiLorenzo, I Gomes, T Salomon, EC Sabino, L Capuani, DTS Cruz, C Maximo, MV Flor-Park, RA Mota, DO Werneck Rodrigues, CL Dinardo, C Almeida-Neto, B Custer, S Kelly, Recipient Epidemiology and Donor Evaluation Study-III (REDS-III) International Component Brazil

**Author notes:** **Corresponding Author:** Shannon Kelly, MD, PhD, Email adress.

## Abstract

**Introduction:** Many individuals with sickle cell disease (SCD) die before age 60, despite early detection via neonatal screening and implementation of treatments such as vaccines and antibiotic prophylaxis and the increasing availability of disease modifying therapies. This study evaluated the causes and independent predictors of mortality in a SCD population in Brazil.

**Methods:** This analysis was performed within the multicenter Recipient Epidemiology and Donor Evaluation (REDS)-III SCD cohort which was established at 6 participating centers in Brazil from 2013-2018. Participants were randomly selected as eligible and recruited at routine visits. Medical records were reviewed to abstract clinical and laboratory data. Mortality and cause of death were confirmed by local chart review as well as linkage to the Brazilian death certificate database. Key variables were compared between deceased and alive participants using Chi^2^ test for categorical variables and Mann-Whitney test for continuous variables. Stepwise logistic regression then a Cox regression multivariable model was performed to identify independent predictors for mortality within the adult participants.

**Results:** There were 2,793 participants in the cohort (1,558, 55.8%, <18 years) and 159 (5.7%) were confirmed to be deceased by the end of follow up: 142 adults (>18 years) and 17 children. The median life expectancy was 65.7. Within adults, infection was the main identifiable cause of death (33.3%), followed by pulmonary causes (25.2%) and neurologic causes (14.5%). Five (3.1%) patients had an unknown cause of death. Independent predictors of mortality were age [Hazard Ratio (HR) 1.03; 95% CI 1.01-1.04; p<.01], iron overload (HR 1.68; 95% CI1.09-2.60; p<.02] and previous hospital admission (HR 1.68; 95% CI 1.10-2.56; p<.02).

**Discussion:** Mortality in Brazilian SCD individuals is shifting from children to adults, with increased rates of death in the third and fourth decades of life. Individuals with SCD are dying 10 years before the general population in Brazil. The main causes of death in our cohort were infections, acute chest syndrome and stroke, highlighting the need for prompt recognition and treatment of these complications. Screening and treatment for iron overload and closer monitoring and consideration of disease modifying therapies for patients with frequent hospital admissions are important as both were identified as independent predictors of mortality.

## INTRODUCTION

Most individuals with sickle cell disease (SCD) die before age 60[1–3]. Early diagnosis via neonatal screening, vaccines and antibiotic prophylaxis can decrease infectious complications and childhood mortality[2]. In countries where these preventive measures are commonly implemented, more than 95% of children survive into adulthood[4]. However, adults with SCD suffer from chronic complications that may be less responsive to treatment, despite disease modifying therapies such as hydroxyurea or chronic red blood cell transfusions. The only curative treatment currently available is hematopoietic stem cell transplantation (HSCT), which may not be an option for all people with SCD[5]due to the lack of matched sibling donors, access to a medical center that provides HSCT, and the financial cost. HSCT with alternative donors, such as unrelated or haploidentical donors, and other promising new technologies, such as gene therapy and gene editing, are currently still primarily performed in the setting of a clinical trial.

Survival estimates for people with SCD vary significantly worldwide. In the United States, the National Institutes of Health Cooperative Study of SCD estimated a median survival of 42 years for SCD men and 48 years for women in 1994[1]. In 2016, Gardner et al. published their experience at a single center in the United Kingdom (UK) from 2004-2013 that revealed a median survival estimate of 67 years for SCD individuals[6] as compared to 80.3 years for men and 84.2 for women in the UK. This longer survival estimate, however, reflects the quality of care and treatment in a high-income country. In Brazil, a middle-income country with inequities in access to health care[7], people with SCD face an increased mortality rate at a younger age. Lobo et al. reported a 53.3-year-old life-expectancy after 15-years of follow-up of a SCD cohort in Rio de Janeiro, Brazil in 2018[8]. Life-expectancy for the general population in Brazil was 74.6 years at the time[9]. A metanalysis of eight studies carried out in five states in Brazil published in 2017[10] described high rates of death in SCD populations within different regions of Brazil. This ranged from 3.7% (10-year follow-up period) to 7.4% in children (14-year period of follow-up) and up to 15.9% in adolescents/adults (30-year follow-up period). This mortality in SCD children is much higher than the general mortality rate in the pediatric population in Brazil. UNICEF published the most recent mortality data for Brazilian children under 5-years-old in 2020, reporting mortality in 1.2% of females and 1.6% of males[11]

SCD has major impacts on the quality of life and life-expectancy of affected individuals in Brazil. A comprehensive understanding of mortality for SCD is needed for health care professionals to identify the patients at highest risk of death and then target treatments and preventative strategies to improve their care. To address this need, we conducted a study to identify the mortality rate and independent predictors of mortality of participants enrolled into the Recipient Epidemiology and Donor Evaluation Study-III (REDS-III) SCD cohort at six blood centers in Brazil from 2013 to 2018.

## METHODS

### 1. The multicenter Recipient Epidemiology and Donor Evaluation Study-III (REDS-III) program

This cohort was part of the multicenter REDS-III program funded by the National Heart, Lung, and Blood Institute (NHLBI) of the USA National Institutes of Health. The REDS-III program conducted research focused on blood safety and availability, and the impact of transfusion in recipients, in the USA, Brazil, China, and South Africa[12]. The REDS-III Brazil SCD cohort study was designed to investigate the impact of transfusion on disease outcomes and was a collaboration between Vitalant Research Institute (VRI) in San Francisco, CA, and participating blood centers in Brazil. Research Triangle Institute, International (RTI), served as the data coordinating center. Details of the cohort study procedures and enrollment findings have been described previously[13].

### 2. Study setting

The participating blood centers were Hemope, located in the northeast of Brazil; and five centers in the southeast of Brazil including Hemorio in Rio de Janeiro, Hemominas in the cities of Belo Horizonte (BH), Montes Claros (MC) and Juiz de Fora (JF); and the Instituto da Criança-Hospital das Clínicas da Faculdade de Medicina da Universidade de São Paulo (HC) in São Paulo.

### 3. Study design and population

The six participating centers collectively treat almost 10,000 SCD patients. A random list of almost half (i.e., 4,956 of 9,676) of the patients with at least 1 clinical encounter in the past 3 years was generated to define eligible participants in strata proportional to each center’s SCD population, based on distribution of gender, age (children <18 years and adults ≥18 years) and SCD genotype. The 4956 individuals were selected to oversample enough participants to achieve a recruitment goal of 3,000 participants. SCD patients treated at the six centers were invited to participate in the cohort study at routine patient care visits, and signed informed consent if they agreed to participate.

### 4. Study procedures

At enrollment, an interview was performed to collect data related to demographics, current work/school and marital status, education level, monthly income, and health insurance. Participants were questioned by trained research staff in a private setting to maintain confidentiality. Medical records were reviewed by physicians or nurses/biologists trained by physicians who abstracted clinical and laboratory data based on standardized definitions[13]. For example, the definition of iron overload was ferritin > 1000ng/mL and having received ≥ 20 red blood cell unit lifetime transfusions according to these published definitions. Two follow-up visits occurred approximately annually to conduct updated interviews and abstract new clinical events from the medical record data. A centralized database was created to store collected data. Blood samples were also collected to create a biorepository and perform routine laboratory testing.

### 5. Confirmation of death status

The primary outcome in this analysis was mortality. To confirm death status, a multi-layer approach was implemented. Participant status at the end of the follow-up period was confirmed locally at each center but also by linkage with the Brazilian Ministry of Health death certificate database. Due to the absence of a national identity number in Brazil, a two-step linkage procedure was used. First, an in-house linkage algorithm was developed by staff members from the Brazilian Ministry of Health, using only the person’s name and date of birth to link cohort participants to individuals identified in the death certificate database. Because high sensitivity was achieved in this step at the expense of low specificity, a second step was necessary to increase the specificity of the matches found in the first step. REDS-III study researchers used the Reclink III open-source software to link cohort participants to the Ministry of Health death certificate from 2000 – 2017 using mother’s name in addition to patients’ name and date of birth as the matching variables. This linkage procedure has been validated in a previous publication, in which linkage results were compared from blood donors whose vital status was known, and the results indicated a sensitivity of 94% (95%CI, 90–97%) and a specificity of 100% (95%CI, 98%-100%) for confirmation of death status[14].

Cause of death was determined by a combination of data obtained from ICD-10 codes from the death certificate, but also by reviewing the cause of death entered in the study database by REDS investigators. In some cases, only one source was available and, therefore, it was used to define the cause of death. If both sources were available and there was a discrepancy between the two, a panel of hematologists with expertise in SCD reviewed the case to determine a cause of death.

### 6. Statistical analysis

Clinical and laboratory variables were compared between those who died or survived in an analysis stratified by age (<18 years and ≥18 years). Categorical variables such as demographics, sickle cell genotype, any hospital admission during REDS-III, presence of clinical complications, and treatments during the follow-up period were compared using the Chi^2^ test. Continuous variables, such as laboratory results, were calculated as a mean of results from the three visits (enrollment, follow-up 1 and 2) and the difference analyzed by the Mann-Whitney test.

Only adults ≥18 years were included in the multivariable analysis, as we had a small number of deaths within participants younger than 18 years old. This was performed in two steps. First, all clinical and laboratory variables with a p<0.20 associated with death in the initial bivariate analysis were included in a stepwise logistic regression model. This model selected variables independently associated with death to then be included in a final multivariable Cox regression model.

The method used to calculate the median life expectation was the Kaplan-Meier product-limit estimator. This non-parametric technique estimates the survival probability as a function of time for defined time intervals. The person-time was calculated by the sum of time each participant remained in the study before dying or the final period of observation (i.e., 1808 days). We calculated the number of deaths by person-time to obtain the incidence (for death) rate. We considered the 50^th^ percentile of survival time to the life-expectancy of the general population. Stata ^®^17.0 software was used to perform all statistical analysis.

### 7. Ethical considerations

This study was approved by all six blood centers’ ethical committees, the Brazilian National Ethical Committee for Research (CAAE: 46981615.0.1001.0065), and the Institutional Review Board at Vitalant Research Institute (VRI) in San Francisco, CA. All participants that agreed to participate in the study signed a written informed consent.

## RESULTS

There were 2,793 participants enrolled into the cohort from 2013-2015 and followed through 2018. A little over half of the participants (55.8%, 1,558) were younger than 18 years old upon enrollment (Table 1). There were 159 (5.7%) participants who died during the follow-up period [142 adults and 17 children (Table 2)]. The total person-time was 62,235 days/ year. The death incidence rate was 2.5/1000 person-days The median life expectancy was 65.7 years and percentage of death was similar among females (6.2%) and males (5.1%). Most pediatric patients died during adolescence (11-17 years old) (47% of deceased children or 5% of all deceased patients) while most deceased adult patients were older than 36 years old (52.8%). About half of the adult patients who were alive at end of study were working or in school at last follow-up, as compared with only 40.8% of those who had died by the end of the study (p=0.03).

**Table 1.**
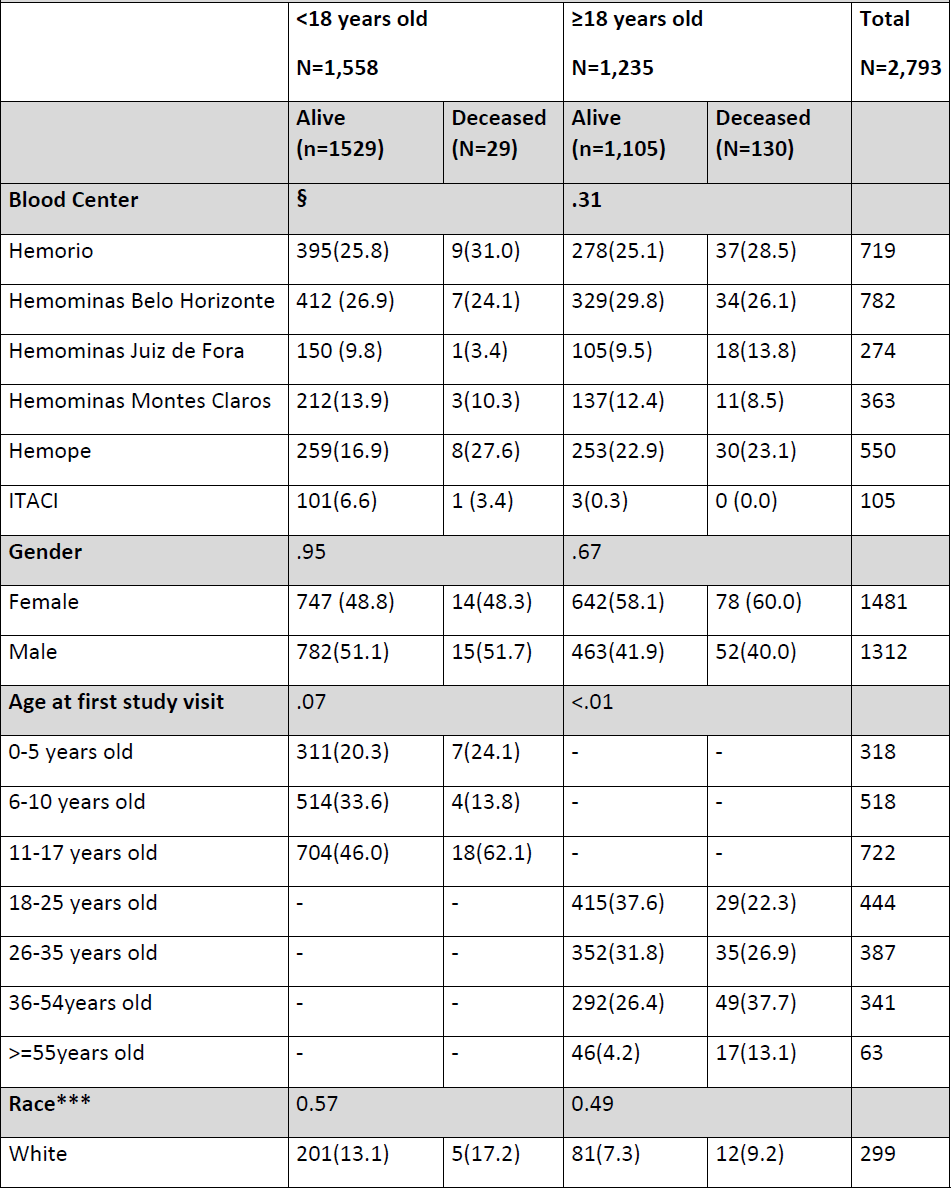
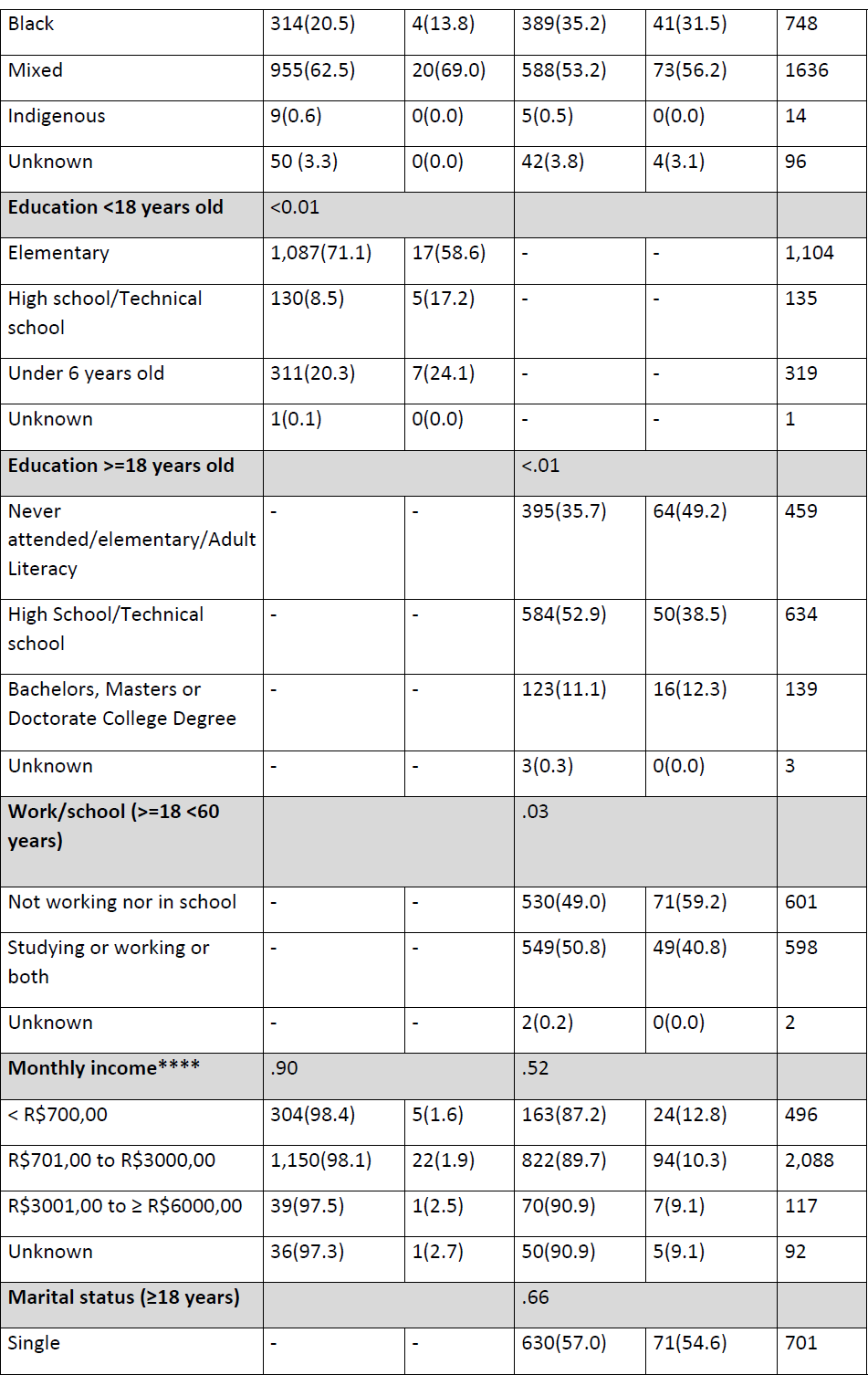
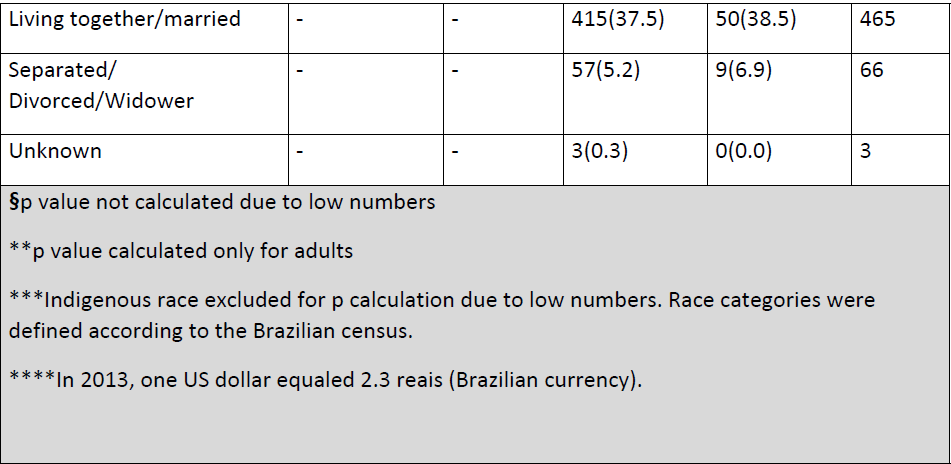
Demographic data of REDS-III Adults and Children According to Vital Status.

|  |  |  |  |  |  |
| --- | --- | --- | --- | --- | --- |
|  | <18 years old<br>N=1,558 |  | ≥18 years old<br>N=1,235 |  | Total<br>N=2,793 |
|  | Alive<br>(n=1529) | Deceased<br>(N=29) | Alive<br>(n=1,105) | Deceased<br>(N=130) |  |
| **Blood Center** | § |  | .31 |  |  |
| Hemorio | 395(25.8) | 9(31.0) | 278(25.1) | 37(28.5) | 719 |
| Hemominas Belo Horizonte | 412 (26.9) | 7(24.1) | 329(29.8) | 34(26.1) | 782 |
| Hemominas Juiz de Fora | 150 (9.8) | 1(3.4) | 105(9.5) | 18(13.8) | 274 |
| Hemominas Montes Claros | 212(13.9) | 3(10.3) | 137(12.4) | 11(8.5) | 363 |
| Hemope | 259(16.9) | 8(27.6) | 253(22.9) | 30(23.1) | 550 |
| ITACI | 101(6.6) | 1 (3.4) | 3(0.3) | 0 (0.0) | 105 |
| **Gender** | .95 |  | .67 |  |  |
| Female | 747 (48.8) | 14(48.3) | 642(58.1) | 78 (60.0) | 1481 |
| Male | 782(51.1) | 15(51.7) | 463(41.9) | 52(40.0) | 1312 |
| **Age at first study visit** | .07 |  | <.01 |  |  |
| 0-5 years old | 311(20.3) | 7(24.1) | - | - | 318 |
| 6-10 years old | 514(33.6) | 4(13.8) | - | - | 518 |
| 11-17 years old | 704(46.0) | 18(62.1) | - | - | 722 |
| 18-25 years old | - | - | 415(37.6) | 29(22.3) | 444 |
| 26-35 years old | - | - | 352(31.8) | 35(26.9) | 387 |
| 36-54years old | - | - | 292(26.4) | 49(37.7) | 341 |
| >=55years old | - | - | 46(4.2) | 17(13.1) | 63 |
| **Race\*\*\*** | 0.57 |  | 0.49 |  |  |
| White | 201(13.1) | 5(17.2) | 81(7.3) | 12(9.2) | 299 |
| Black | 314(20.5) | 4(13.8) | 389(35.2) | 41(31.5) | 748 |
| Mixed | 955(62.5) | 20(69.0) | 588(53.2) | 73(56.2) | 1636 |
| Indigenous | 9(0.6) | 0(0.0) | 5(0.5) | 0(0.0) | 14 |
| Unknown | 50 (3.3) | 0(0.0) | 42(3.8) | 4(3.1) | 96 |
| **Education <18 years old** | <0.01 |  |  |  |  |
| Elementary | 1,087(71.1) | 17(58.6) | - | - | 1,104 |
| High school/Technical school | 130(8.5) | 5(17.2) | - | - | 135 |
| Under 6 years old | 311(20.3) | 7(24.1) | - | - | 319 |
| Unknown | 1(0.1) | 0(0.0) | - | - | 1 |
| **Education >=18 years old** |  |  | <.01 |  |  |
| Never attended/elementary/Adult Literacy | - | - | 395(35.7) | 64(49.2) | 459 |
| High School/Technical school | - | - | 584(52.9) | 50(38.5) | 634 |
| Bachelors, Masters or Doctorate College Degree | - | - | 123(11.1) | 16(12.3) | 139 |
| Unknown | - | - | 3(0.3) | 0(0.0) | 3 |
| **Work/school (>=18 <60 years)** |  |  | .03 |  |  |
| Not working nor in school | - | - | 530(49.0) | 71(59.2) | 601 |
| Studying or working or both | - | - | 549(50.8) | 49(40.8) | 598 |
| Unknown | - | - | 2(0.2) | 0(0.0) | 2 |
| **Monthly income\*\*\*\*** | .90 |  | .52 |  |  |
| < R\$700,00 | 304(98.4) | 5(1.6) | 163(87.2) | 24(12.8) | 496 |
| R\$701,00 to R\$3000,00 | 1,150(98.1) | 22(1.9) | 822(89.7) | 94(10.3) | 2,088 |
| R\$3001,00 to ≥ R\$6000,00 | 39(97.5) | 1(2.5) | 70(90.9) | 7(9.1) | 117 |
| Unknown | 36(97.3) | 1(2.7) | 50(90.9) | 5(9.1) | 92 |
| **Marital status (≥18 years)** |  |  | .66 |  |  |
| Single | - | - | 630(57.0) | 71(54.6) | 701 |
| Living together/married | - | - | 415(37.5) | 50(38.5) | 465 |
| Separated/<br>Divorced/Widower | - | - | 57(5.2) | 9(6.9) | 66 |
| Unknown | - | - | 3(0.3) | 0(0.0) | 3 |
| §p value not calculated due to low numbers<br>**p value calculated only for adults<br>***Indigenous race excluded for p calculation due to low numbers. Race categories were defined according to the Brazilian census.<br>****In 2013, one US dollar equaled 2.3 reais (Brazilian currency). |  |  |  |  |  |

**Table 2.**
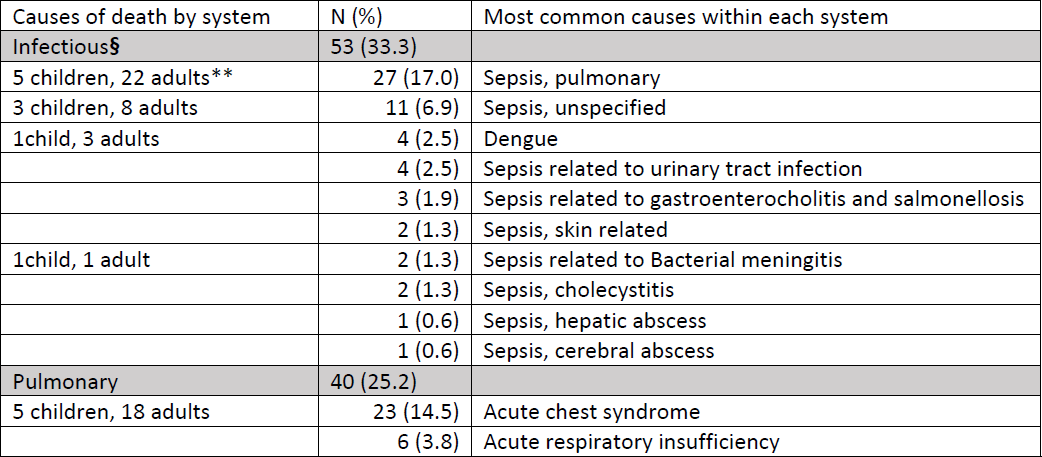
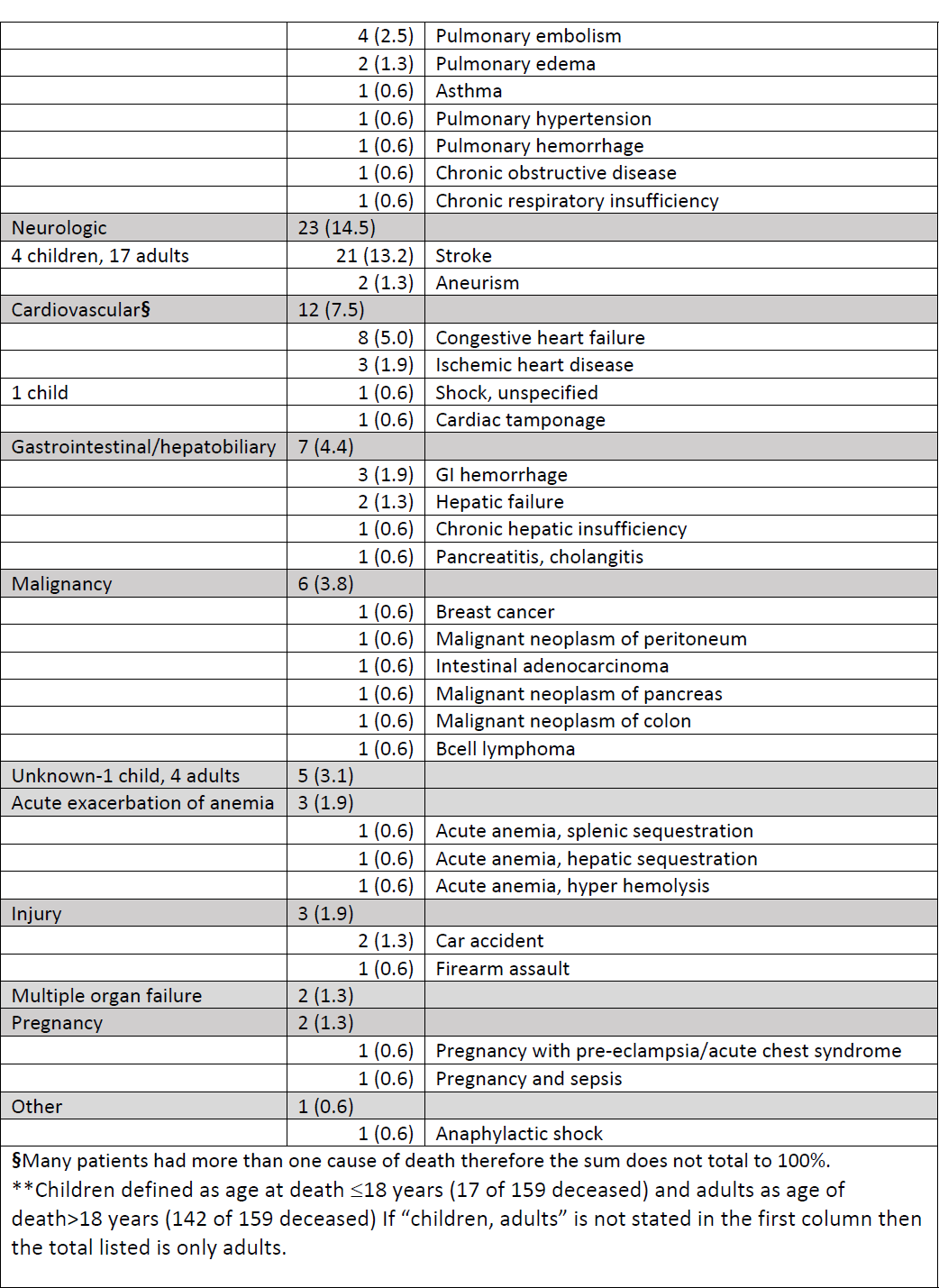
Causes of death among 159 Deceased Participants in the REDS-III Brazil SCD Cohort.

Table 2 shows the causes of death among our deceased participants. Infection was the most common identifiable cause of death (33.3%), followed by pulmonary (25.2%) and neurologic (14.5%) causes. In detail, sepsis secondary to pulmonary infection was the main cause of death (17%), followed by acute chest syndrome (14.5%) and stroke (13.2%). Five (3.1%) patients had an unknown cause of death.

Comparison of laboratory results of adults showed a lower mean hemoglobin (8.2g/dL vs 9.3g/dL; p<0.01), lower mean platelet count (336,591/mm^3^ vs 363,006/mm^3^; p<0.03) and higher mean creatinine (1.1mg/dL vs 0.74mg/dL; p<0.01) in deceased participants compared to living participants. Reticulocytes and direct bilirubin were higher and fetal hemoglobin lower in deceased patients (not statistically significant, all lab comparisons shown in Table 3). The average number of lifetime red blood cells units transfused was 35 in deceased vs. 22 in living participants (p<0.01).

**Table 3.** Comparison of laboratory values between alive and deceased adults within the REDS-III Brazil SCD Cohort.

| Laboratory Results<br>Mean (SD) /range | Alive (n=1105) | Deceased (N=130) | N | Wilcoxon<br>rank |
| --- | --- | --- | --- | --- |
| Hemoglobin (g/dL) | 9.3(1.9)/ 4.8-15.7 | 8.2(1.9)/ 3.5-13.2 | 1192 | <.01 |
| Leucocytes (cells/ mm<sup>3</sup>) | 10110(3680)/ 3065-30073 | 10143(3703)/ 2770-21100 | 1190 | .93 |
| Platelets (number/ mm<sup>3</sup>) | 363,006(134,465)/ 82,933-786,000 | 336,591(126,844)/ 105,000-663,500 | 1189 | .03 |
| Creatinine (mg/dL) | 0.74(0.35)/ 0.21-6.6 | 1.1(1.1)/ 0.4-9.7 | 1235 | <.01 |
| Reticulocytes (%) | 8.6(4.6)/ 0.3-32.5 | 9.3(5.1)/ 1-28 | 1126 | .18 |
| Direct Bilirubin (mg/dL) | 0.6(1.5)/ 0.01-45 | 0.74(0.8)/ 0.02-6.5 | 1235 | .10 |
| Fetal Hemoglobin (%) | 10.7(9.1)/ 0.1-47 | 8.8(8.5)/ 0.6-36 | 694 | .13 |
| Bilirubin total | 1.9(1.6)/ 0.14-17.7 | 2(1.7)/ 0.2-11.8 | 1235 | .57 |
| Average number of RBC<br>units transfused during<br>whole life (mean) | 22.2(33.4) | 35.3(40.4) | 1108 | <.01 |
| Serological Status<br>(positive) | Alive (n=1105) | Deceased (N=130) | Not<br>Tested | Chi 2 |
| Syphilis (VDRL) | 28 (2.5) | 5 (3.8) | 3(0.25) | .38 |
| HBV (HBsAg) | 9(0.8) | 2 (1.5) | 1(0.1) | .66 |
| HCV (anti-HCV) | 55 (5.0) | 13 (10.0) | 1 (0.1) | .05 |
| HIV (anti-HIV 1/2) | 5(0.4) | 2 (1.5) | 2 (0.2) | .26 |
| HTLV (anti-HTLV-1/2) | 15 (1.4) | 1 (0.8) | 2 (0.2) | .83 |
| Chagas (anti-Chagas) | 10 (0.9) | 3 (2.3) | 2 (0.2) | .09 |
| Negative for all tests | 989(89.5) | 105(80.8) | 3(0.25) | --- |

Table 4 details clinical predictors associated with mortality in adults in bivariate analysis. Deceased patients were more likely to have systemic arterial hypertension (36.9% vs. 14.4%; p<.01), iron overload (31.5% vs. 16.6%; p<.01), leg ulcers (26.1% vs. 18.3%; p<.03), pulmonary hypertension (24.4% vs. 9.8%; p<.01), delayed hemolytic transfusion reaction (3.1% vs. 0.9%; p<.03), pancreatitis (3.1% vs. 0,9%; p<.03), venous thromboembolism (6.9% vs. 2.6%; p<.01) and sepsis (11.5% vs. 6.7%; p<.04). Hospital admissions during the past year were also more frequent among deceased patients (70.8% vs. 49.1%; p<.01).

**Table 4.** Clinical predictors associated with Mortality Among Adults in the REDS-III Brazil SCD Cohort.

| Variables | Alive (n=1105) | Deceased (n=130) | Chi 2(p value) |
| --- | --- | --- | --- |
| **Sickle Cell Disease Genotype** |  |  |  |
| SS | 801(88.4) | 105(11.6) | .18 |
| SC | 229(92.7) | 18(7.3) |  |
| Sβ<sup>0</sup>Thalassemia (Sβ<sup>0</sup>) | 37(90.2) | 4(9.8) |  |
| Sβ<sup>+</sup>Thalassemia (Sβ<sup>+</sup>) | 31(91.2) | 3(8.8) |  |
| Other | 7(100) | 0(0.0) |  |
| **Clinical Complications** |  |  |  |
| Systemic arterial hypertension | 159(14.4) | 48(36.9) | <.01 |
| Iron overload | 183(16.6) | 41(31.5) | <.01 |
| Leg ulcers | 202(18.3) | 34(26.1) | .03 |
| Pulmonary hypertension | 108(9.8) | 30(24.4) | <.01 |
| Delayed hemolytic transfusion reaction | 10(0.9) | 4(3.1) | .03 |
| Pancreatitis | 10(0.9) | 4(3.1) | .03 |
| Venous thromboembolism | 29(2.6) | 9(6.9) | <.01 |
| Sepsis | 74(6.7) | 15(11.5) | .04 |
| Hospital admissions during past year | 543 (49.1) | 92(70.8) | <.01 |
| **Medical Treatments** |  |  |  |
| Chronic Red Blood Cells Transfusion | 111(10.1) | 23(17.7) | <.01 |
| Analgesics (>30days) | 205(18.6) | 34(26.1) | .04 |
| Hydroxyurea | 567(51.3) | 72(55.4) | .38 |
| Antidepressants | 133(12.1) | 18(13.9) | .55 |
| The following clinical predictors were not significantly different when compared between alive and deceased patients: ischemic stroke, neuropathic pain, pulmonary embolism, cholecystitis, cholelithiasis, hepatic and splenic sequestration, any previous vaso-occlusive pain, previous surgery, splenectomy, cholecystectomy. |  |  |  |

A comparison of treatments between those alive and deceased is summarized in Table 4. Chronic transfusions (17.7% vs. 10.1%; p<.01) and the use of chronic pain medication (26.1% vs. 18.6%; p<.04) were more frequent among the deceased. Hydroxyurea was used by half of participants in both groups and antidepressant use was also not different between the two groups.

Table 5 shows the independent predictors associated with mortality within adults identified by Cox logistic regression. Older age showed a significant trend towards increased odds of death. Iron overload increased the risk of death by more than 1.5 times (95% IC 1.09-2.60; p<.02). Any previous hospital admissions also increased the risk of death by more than 1.5 times (95% IC 1.10-2.56; p<.02).

**Table 5.** Multivariable cox regression analysis of predictors associated with mortality in SCD adult patients in Brazil (2013-2018)

|  | Hazard Ratio | Confidence interval | Pvalue |
| --- | --- | --- | --- |
| Age (continuous) | **1.03** | **1.01-1.04** | **.01** |
| Creatinine | 1.09 | 0.93-1.27 | .27 |
| Hemoglobin interval range§ |  |  |  |
| >9.14 (Reference) | 1 | - | - |
| 8.17-9.14 | 1,36 | 0.82-2.27 | .24 |
| <8.16 | 1,16 | 0.67-2.00 | .60 |
| Iron overload | **1.68** | **1.09-2.60** | **.02** |
| Previous hospital admission | **1.68** | **1.10-2.56** | **.02** |
| Pancreatitis | 2.63 | 0.91-7.63 | .07 |
| Pulmonary hypertension | 1.40 | 0.88-2.25 | .16 |
| Systemic Arterial Hypertension | 1.26 | 0.79-2.01 | .33 |
| Sepsis | .74 | 0.39-1.40 | .36 |
| Cholecystitis | 1.19 | 0.74-1.89 | .47 |
| Hydroxyurea use** | 0.96 | 0.65-1.42 | .84 |

Stepwise cox logistic regression was applied in a multivariable model including all clinical and laboratory variables with a $p < 0.05$ in bivariate analysis. Hazard ratio value is compared to “no” category not shown for each variable, except for continuous variables.
§Hemoglobin interval range was defined by distribution in tertiles.
\*\*Hydroxyurea was included in analysis due to its important ability to modify clinical and laboratory variables despite $p > 0.05$ in the initial analysis

## DISCUSSION

Death due to SCD remains a substantial health problem in Brazil. During the 5-year-period of this cohort study, our patients showed a lower median life expectancy when compared to the general population in Brazil (65.7 years vs 76.7 years)[15]. Our cohort demonstrated a higher death rate among adults in their third and fourth decades of life, but decreasing rates of death among children as compared to historical controls, reflecting a shift from early mortality to late mortality in adult patients. Our data are similar to a prospective study from Tanzania[16], that also reported 5.7% of their cohort had died, although the African study demonstrated a higher rate of death among children. We presume this difference reflects improved access to vaccines and antibiotic prophylaxis in Brazil. Other studies from the US also demonstrated a similar trend of increased death rate among adults when compared to children[17, 18], which may reflect the difficulties in finding appropriate health care and treatment for adult SCD patients with chronic complications [18].

Our study was able to determine many of the specific causes of mortality in this Brazilian SCD cohort. The causes of death were mainly related to infections, acute chest syndrome, stroke and cardiovascular disease. Our results are similar to another Brazilian study[8] published in 2018 that demonstrated the most common causes of death in 1,676 patients treated at Hemorio were infection (29.2%), followed by acute chest syndrome (25.3%) and stroke (11.7%). An US cohort study published in 2013 reviewed ICD-10 causes of death among SCD patients and also found infection as the main cause of death in their population[18]. Therefore, despite increased access to vaccines and guidelines for prompt antibiotic treatment, individuals with SCD remain at increased risk for death with infection, even in current times and even in high income countries.

The main clinical predictors of mortality in our cohort were increasing age, iron overload and previous hospital admission. Increasing age is likely related to the progressive deterioration and multiorgan disease and dysfunction that occurs in SCD. Organ dysfunction may not be always clinically recognized but may contribute to increased risk of death, as previous studies highlighted the importance of lung disease, renal failure, and central nervous system deterioration in causing premature death among SCD population compared to non-SCD adults[19].

The identification of iron overload as a significant predictor of death in our cohort is critically important. Acute transfusions treat several complications in SCD such as acute chest syndrome, splenic sequestration, and aplastic crisis, and chronic transfusion therapy can prevent stroke, a severe and devastating complication of SCD. But repeated transfusions also cause accumulation of iron, which must be continuously monitored and treated with iron chelation therapy when iron overload develops. The mean number of transfused RBC units among deceased participants was almost double that transfused to participants confirmed to be alive by the end of follow up. While frequent transfusions may be an indicator of severe disease, iron overload - not the number of transfusions - was an independent predictor or mortality in our data. Iron overload is known to be associated with liver disease in patients with SCD[20–22], and well recognized to cause higher mortality in chronically transfused populations such as thalassemia patients[20]. The 2020 American Society of Hematology guidelines for SCD support recommended screening chronically transfused SCD patients with magnetic resonance imaging (MRI) to measure liver iron content every 1-2 years compared to screening with ferritin alone [23]. These guidelines do not address screening guidelines for patients not treated with chronic transfusion therapy, yet some patients with severe disease receive many acute transfusions and are also at risk for iron overload. Therefore, guidelines might consider addressing screening for iron overload for patients not treated with chronic transfusion therapy yet having received ≥ 20 red blood cells units during their lifetime. For example, our data showed that among the 130 adults who died, although 88 (67.7%) did not have iron overload and never received chronic transfusion therapy, 18 (13.8%) had iron overload and received chronic transfusion therapy, and, importantly, 23 (17.7%) had iron overload but never received chronic transfusion therapy. There was only 1 person (0.8%) in the deceased group who received chronic transfusion therapy but did not have iron overload.

One limitation of our analysis is the use of ferritin to define iron overload rather than a more specific measure of tissue iron by MRI or other techniques. We chose this definition as ferritin was measured in the entire cohort in a standardized way as part of this research study and is commonly used for iron screening in Brazil where not all sites have access to MRI for iron measurement[24]. Most guidelines recommend treatment with iron chelation once patients demonstrate a serum ferritin >1000 μg/l, or Liver Iron Concentration > 3 mg/g dry weight liver[23, 25]. This is the standard treatment in Brazil and oral chelation treatment is provided free of charge by the universal health system (SUS)[26]. However, despite the free treatment in Brazil, the main obstacles to iron overload treatment remains the difficult adherence to a chronic medication[25] as well as underdiagnosis due to lack of access to MRI and ferritin tests in some locations managed by the public health system. Most of our deceased cohort participants with iron overload as a diagnosis had acute chest syndrome as the cause of death, followed by sepsis, cardiac causes, and stroke, rather than a liver specific diagnosis as a cause of death. Increased iron is known to increase the risk of infection and may also increase the risk of other complications, or work synergistically to increase morbidity/mortality associated with those complications. Further research is warranted to elucidate the mechanisms that contribute to increased odds of death in individuals with SCD and iron overload.

The third independent predictor of mortality identified in our analysis, a history of previous hospital admission, increased the chance of death by 50% in our participants. We were not able to analyze the time interval between a hospital admission and death. It is possible that a history of hospital admission is a surrogate marker of disease severity. Alternatively, hospital admissions may directly contribute to the cause of death. For example, Fung et al.[20] demonstrated that among 199 American patients with SCD, pain was the most common cause for hospitalization and pain may act as a precursor to severe and life-threatening complications as acute chest syndrome and sudden unexpected death[5]. Individuals with SCD with history of hospitalization, particularly those with frequent hospitalizations, warrant more frequent follow-up and consideration for disease modifying or curative therapies.

We found that homozygous HbSS was the predominant genotype associated with death, as previously described by Platt in 1994[1], confirming the increased severity of this genotype as compared to others. However, we did not find differences in mortality between those participants treated, or not, with hydroxyurea. This lack of association may be due to the shorter time of follow-up in our cohort (5 years) versus other studies that showed improved survival among hydroxyurea users[8, 27]. In addition, our study was not designed to evaluate the impact of this treatment on mortality, as we did not collect specific data on adherence and timeline of medication use. The Platt et al. study also identified that higher leukocyte counts and lower fetal hemoglobin were associated with increased mortality in SCD[1]. While these were not independent predictors of mortality in our cohort, it is possible that the increasing use of hydroxyurea between the time of that study and ours may have altered the relationship between these laboratory values and mortality.

Our study has limitations. Clinical and laboratory data were abstracted from medical records. While we used standardized definitions to abstract data and performed multiple levels of quality control assessment and verification, it is possible that missing data or errors in the abstraction process occurred. In terms of the generalizability of our results, our six participaing sites do not include all regions in Brazil. However we do have sites in the locations where most patients are treated. We studied mortality in our adult population only because we, fortunately, had few deaths among our pediatric participants, therefore we were not able to analyze the predictors of death among children. Pulmonary hypertension, a major predictor for death in SCD adults according to other studies[18], was not recognized as a predictor for mortality in our cohort. We believe this lack of association was secondary to the difficulty in accessing echocardiograms at most of our participating sites and, therefore, led to the underdiagnosis of this clinical complication in our cohort.

## CONCLUSIONS

We believe our study sheds light on the causes and predictors of mortality among the sickle cell disease population in Brazil as a whole, and our findings may contribute to the development of strategies to increase survival rates among this patient population. We recommend early recognition of iron overload, including consideration of screening in patients not treated with chronic transfusion therapy who have received >20 red blood cell units for acute indications. Emphasis should be placed on adherence to iron chelation therapy for all patients with iron overload in addition to closer monitoring of patients with frequent hospitalization who should be considered for disease modifying therapies.

Mortality is shifting from pediatric patients to adults, most likely due to improved survival after early recognition of disease in newborns, vaccination and prophylactic antibiotics. Yet infection remained a common cause of death in this current day cohort, emphasizing the importance of prompt evaluation of all SCD individuals with fever and urgent management of suspected sepsis.

Individuals with SCD in Brazil are dying 10 years before the general population. Training health professionals in recognizing the association of these conditions and death in patients with sickle cell disease is critical and ensuring easy access to monitoring and treatment is urgent for improving survival for this vulnerable population.

## Data Availability

Authors are able to make fully available and without restriction all data underlying their findings.

## Acknowledgement

Funding for this study was provided by the REDS-III contract of the NHLBI/NIH under HHSN2682011-00007I.

